# Serum neurofilament light as a biomarker for prognosis in patients with newly diagnosed Parkinson’s disease

**DOI:** 10.1101/2021.11.01.21265751

**Authors:** Nirosen Vijiaratnam, Michael Lawton, Amanda Heslegrave, Tong Guo, Manuela MX Tan, Edwin Jabbari, Raquel Real, John Woodside, Katherine A Grosset, Viorica Chelban, Dilan Athauda, Christine Girges, Roger A Barker, John Hardy, Nicholas Wood, Henry Houlden, Nigel M Williams, Yoav Ben-Shlomo, Henrik Zetterberg, Donald G Grosset, Thomas Foltynie, Huw R Morris, PRoBaND clinical consortium

## Abstract

**Background:** Patients with Parkinson’s disease (PD) have variable disease progression. More accurate prediction of progression could improve clinical trial design. Although some variance in clinical progression can be predicted by age at onset and phenotype, we hypothesise that this can be improved by blood biomarkers.

**Objective:** To determine if serum neurofilament light (NfL) is a useful biomarker for prognostic modelling in PD.

**Methods:** We evaluated the relationship between serum NfL and baseline and longitudinal clinical measures as well as patients’ genetic (*GBA* and *APOE*) status in a large clinical dataset. We classified patients as having a favourable or an unfavourable outcome based on a previously validated model and explored whether serum NfL could distinguish prognostic phenotypes. We compared NfL with baseline candidate predictor variables and studied the combination of clinical, serum and genetic data.

**Results:** Serum NfL was associated with patients’ cognitive status at baseline. Baseline NfL was associated with the progression of motor and functional impairment and with increased mortality (Survival HR 1.85, CI: 1.03-3.33, p=0.04). Baseline NfL levels predicted unfavourable progression to a similar extent as previously validated clinical predictors (AUC: 0.74 vs 0.78, p=0.22). The combination of clinical, genetic and biomarker data produced a strong predication of unfavourable outcomes as compared to age and gender alone (AUC: 0.71-age/gender vs 0.83-ALL p = 0.0076)

**Conclusions:** Baseline serum NfL provides an objective measure of neurodegeneration in PD patients. Clinical trials of disease-modifying therapies might usefully stratify patients according using clinical, genetic and NfL status at the time of recruitment.

## INTRODUCTION

Parkinson’s disease (PD) is a progressive neurodegenerative disorder characterised by a wide range of motor and non-motor features which result in substantial morbidity [1]. Disease modification to slow the rate of progression remains a key goal in PD [2]. A challenging aspect is the inherently complex nature of PD with substantial clinical heterogeneity in the rate of progression [3–5]. The underlying basis for this variability is poorly understood but may relate to inflammation, cell to cell spread of pathogenic proteins and compensatory mechanisms. Ultimately, this likely relates to genetic variation [6,7]. Understanding these aspects of disease and investigating the reliability of novel, measurable biological markers of PD severity and progression, and their association with patients’ genetic status will likely form a critical aspect in prognostic prediction which will be important for patient selection in future clinical trials.

Neurofilament light (NfL) is a subunit of neurofilaments, which are structural proteins that confer stability to neurons and are expressed abundantly in larger myelinated axons [8]. Low levels of NfL are constantly released from axons in the healthy state [9]. This increases in response to CNS axonal damage due to inflammatory, neurodegenerative, traumatic or vascular injury.[8] The released NfL enters cerebrospinal fluid (CSF) and subsequently blood, [9] thus making peripheral measurement of NfL a potentially useful biomarker of CNS diseases. Despite its lack of specificity, the association of NfL with axonal injury and the amount of neuronal damage means that it may be useful in predicting progression across a range of neurodegenerative diseases.

This concept has recently been explored in Parkinsonian syndromes [10]. NfL levels correlate with more severe motor and cognitive disease burden at diagnosis and during follow-up, while also potentially predicting subtypes and overall survival rates [11–14] The interaction between patients’ genotype and NfL, as well as their combined usefulness in predicting PD progression remains unclear.

We explored this in a large prospectively followed cohort of patients with a recent diagnosis of PD. We defined whether serum NfL levels could distinguish PD patients from controls. We then determined whether baseline NfL levels corresponded with the severity of symptoms soon after diagnosis, and with subsequent disease progression, while also exploring its interaction with the patients’ underlying genetic status. The potential use of NfL in improving clinical progression modelling for use in clinical trial selection was then explored with the overall hypothesis being that serum NfL would aid in distinguishing between patients with a favourable or unfavourable prognosis.

## METHODS

### Participants

PD participants in this study were recruited from the Tracking Parkinson’s study, a large prospective, observational, multicentre project which recruited patients from February 1, 2012, through to May 31, 2014. The study protocol and baseline patient characteristics have previously been published [15]. Briefly, patients with a clinical diagnosis of PD meeting the Queen Square Brain Bank criteria [16] and supportive neuroimaging (when the diagnosis was not firmly established clinically) were enrolled. Patients had to be within 3.5 years of diagnosis at recruitment and drug-naive and treated patients aged 18 to 90 years were eligible. Exclusion criteria were severe comorbid illness that precluded clinic visits, and other degenerative forms of parkinsonism. Patients were excluded from further follow-up if their diagnosis was revised to an alternative condition.

Patients were selected for NfL analysis based on completion of a minimum follow-up of 2.5 years, with available serum samples at baseline for analysis. Further selection criteria were also applied to provide an equal representation of typical PD with a high index of diagnostic certainty (>95%), and cases with atypical clinical features with a lower index of diagnostic certainty (<80%) at their 2.5-year clinical assessment. Healthy control individuals were enrolled from the PROSPECT study, which is an ongoing natural history cohort study of atypical parkinsonian disorders, including a population of healthy control individuals (consisting of 7 UK study sites with initial recruitment from September 1, 2015, through to December 1, 2018). Control participants included a spouse or a friend of the case or volunteers recruited through the Join Dementia Research volunteer registry. The Tracking Parkinson’s study (REC Reference: 11/AL/0163) and PROSPECT (REC Reference: 14/LO/1575) studies have multicentre research ethics committee approvals.

### Clinical assessments

Baseline demographics such as gender, age, and disease duration were recorded. A detailed description of clinical assessments performed in *Tracking Parkinson’s* have previously been published [15]. In this study we included selective motor (Movement Disorders Society Unified Parkinson’s disease rating scale - MDS-UPDRS, Hoehn & Yahr - H&Y, Schwab and England - S&E), cognitive (Montreal cognitive assessment - MoCA, animal semantic fluency score - SF) and quality of life (PDQ-8) measures. All patients had been diagnosed within the preceding 3.5 years of study entry and underwent assessments every 18 months (although there were some interim visits at 6–12-month intervals which collected other information) and data was available up to visit 10 (72 months). Clinicians determined their diagnostic certainty of PD at each visit (0-100%), while also noting clinical features they deemed to be atypical for PD. Patients who received an alternative diagnosis to PD during follow-up or who had a clinician diagnostic certainty of <90% at the last available visit were excluded from our analysis. All-cause mortality was also noted and studied as a relevant outcome.

#### Favourable vs. Unfavourable outcome subgroups

Patients were classified as having Favourable or Unfavourable outcomes based on a previously validated model of progression [17]. A binary outcome measure was created for unfavourable progression PD (U-PD) when patients had postural instability (defined by a H&Y scale score of 3 or higher) or dementia (a MoCA score of less than 17)[18] at the last available assessment, or if they had died during follow-up. Although the premise for grouping was identical to the previously validated model, outcome measures used to determine this varied slightly due to different scales being used in our cohort (e.g. MMSE substituted with MoCA). All other patients were classified as having favourable progression PD (F-PD). Patients already demonstrating U-PD characteristics at baseline were excluded from the progression to U-PD analyses, but were retained in the baseline analysis and the mixed effects regression analysis [17]. The 3 baseline variables (age at baseline, MDS-UPDRS axial score, and animal SF) that were previously identified to predict the development of U-PD [17] were then explored individually and in combination with NfL to compare clinical and biomarker data in predicting progression.

#### Sample collection and measurement

At enrolment, 10 mL of venous blood was collected from each participant in serum separator tubes. Blood samples were centrifuged (2,500g for 15 minutes) within 1 hour of collection. Serum aliquots were stored in cryotubes at −80°C. Serum NfL concentration was measured using the NF-Light Advantage kit on the HD-X Analyzer (Quanterix, Billerica, MA) by researchers who were blinded to the clinical diagnosis, as previously described [19]. Full details are available on protocols.io: https://dx.doi.org/10.17504/protocols.io.bzbep2je [41].

#### Genetic status classification

Molecular genetic analysis techniques for determining patients *APOE* and *GBA* status have previously been described [7,20]. The step-by-step protocol for SNP genotyping and *APOE* genotyping is available on protocols.io: https://dx.doi.org/10.17504/protocols.io.by9ypz7w [42].

As we and others have previously identified, *APOE* ε4 status is known to be a determinant of cognitive progression, thus patients were classified into groups of either being ε4 carriers (homozygous and heterozygous) and non-carriers [7]. Mutations identified and classification approaches for determining GBA prognostic status in the Tracking Parkinson’s study have previously been detailed [20]. A step-by-step protocol for *GBA* genotyping is available on protocols.io: https: dx.doi.org/10.17504/protocols.io.bzd7p29n [43]. Patients in this study were classified into groups where a GBA variant was identified as either being pathogenic in Gaucher disease (GD) and associated with PD in the heterozygous state (L444P (5 cases), p.R463C (1 case), p.R395C (1 case), p.G377S (1 case), p.N370S (1 case) and p.D409H/L444P/A456P/V460V (1case)), or non-synonymous genetic variants that are associated with PD (E326K (10 cases), T369M (7 cases), and p.D140H/p.E326K (1 case). Two cases with variants of unknown significance were excluded from the group analysis (p.M123T, p.R262H).

### Statistical analysis

Descriptive statistics including mean, standard deviation, median, interquartile range, frequencies, and percentages were used to describe demographic and clinical characteristics by groups. Given non-normally distributed data, differences were compared using Kruskal–Wallis tests for continuous data and chi-squared tests for categorical data. A Natural logarithm (Ln) transformation was performed to reduce right skewness for NfL levels as indicated by inspection of residuals.

The diagnostic accuracy of NfL as a predictive marker of being affected with PD as compared to control status was assessed with logistic regression with age and gender as covariates. Receiver operating characteristic (ROC) analysis with area under the curve (AUC) values with 95% confidence interval (CI) was further determined.

Univariate and multivariable (adjusting for age, gender and disease duration) linear regression analysis was performed to investigate the association between baseline NfL levels and clinical measures of PD. The interaction between *GBA* and *APOE* status with NfL was explored with univariate and multivariate linear regression with NfL as the outcome measure and the respective status of the patients being compared to those who were negative for the respective genetic mutations or alleles of interest. Associations between baseline plasma NfL levels and change in motor, cognitive and quality of life outcomes over time (disease duration from diagnosis as the time axis) were then investigated by linear mixed models, adjusted for age at diagnosis and gender. The mixed models had both a random intercept and a random slope. Cox proportional hazards regression was used to investigate whether the baseline NfL level predicted mortality after adjustment for age and gender.

Logistic regression was repeated using previously validated baseline predictive clinical variables (MDS-UPDRS axial score and semantic fluency) individually and in combination with NfL levels, and the patients’ *GBA* and *APOE* status to explore the ability to distinguish predetermined outcome groups (U-PD vs F-PD). The AUC for each combination of variables was statistically compared against NfL, and together with NfL using Delong’s test.

The Youden J index (maximum sensitivity + specificity – 1) was then calculated for all points of the ROC curve and the maximum value of the index was used as a criterion for selecting the optimum NfL cut-off point for a potential diagnostic test for PD vs controls; and U-PD vs F-PD. All tests were two-sided. All statistical analysis and figures were generated using Stata V.16.1.

## Data and code availability

The original data used in this study is available from the Tracking Parkinson’s (www.trackingparkinsons.org.uk) team. The analysis protocol and code are available at GitHub (https://github.com/huw-morris-lab/proband-nfl) and Zenodo (doi: https://doi.org/10.5281/zenodo.5525370)

### Clinical studies

Tracking Parkinson’s has multi-centre research ethics approval West of Scotland Research Ethics Committee: IRAS 70980, MREC 11/AL/0163, Clinicaltrials.gov: NCT02881099). Each subject provided written informed consent for participation.

## RESULTS

Of the 2000 patients enrolled into the Tracking Parkinson’s study, 291 were studied based on selection criteria. The demographic (age, gender, disease duration from diagnosis) and baseline clinical characteristics (UPDRS 3, H&Y & MOCA) of this cohort was similar to the remaining cohort (Supplementary Table 1). The purpose of this selection approach was to provide good representation of a subset of cases to model progression and to explore the possible use of baseline NfL to determine conversion to an atypical parkinsonian syndrome, in an early Parkinsonism cohort. The number of re-diagnosed cases was low: including 3 cases of progressive supranuclear palsy (PSP), 1 multiple system atrophy (MSA) and 5 with other diagnosis (1 post-polio syndrome, 1 vascular parkinsonism, 1 parkinsonism with a scan without evidence of dopaminergic deficit, 1 essential tremor and 1 uncertain diagnosis). 258 patients were then analysed for progression and phenotype after further exclusion criteria were applied. (Figure 1).

**Figure 1.**
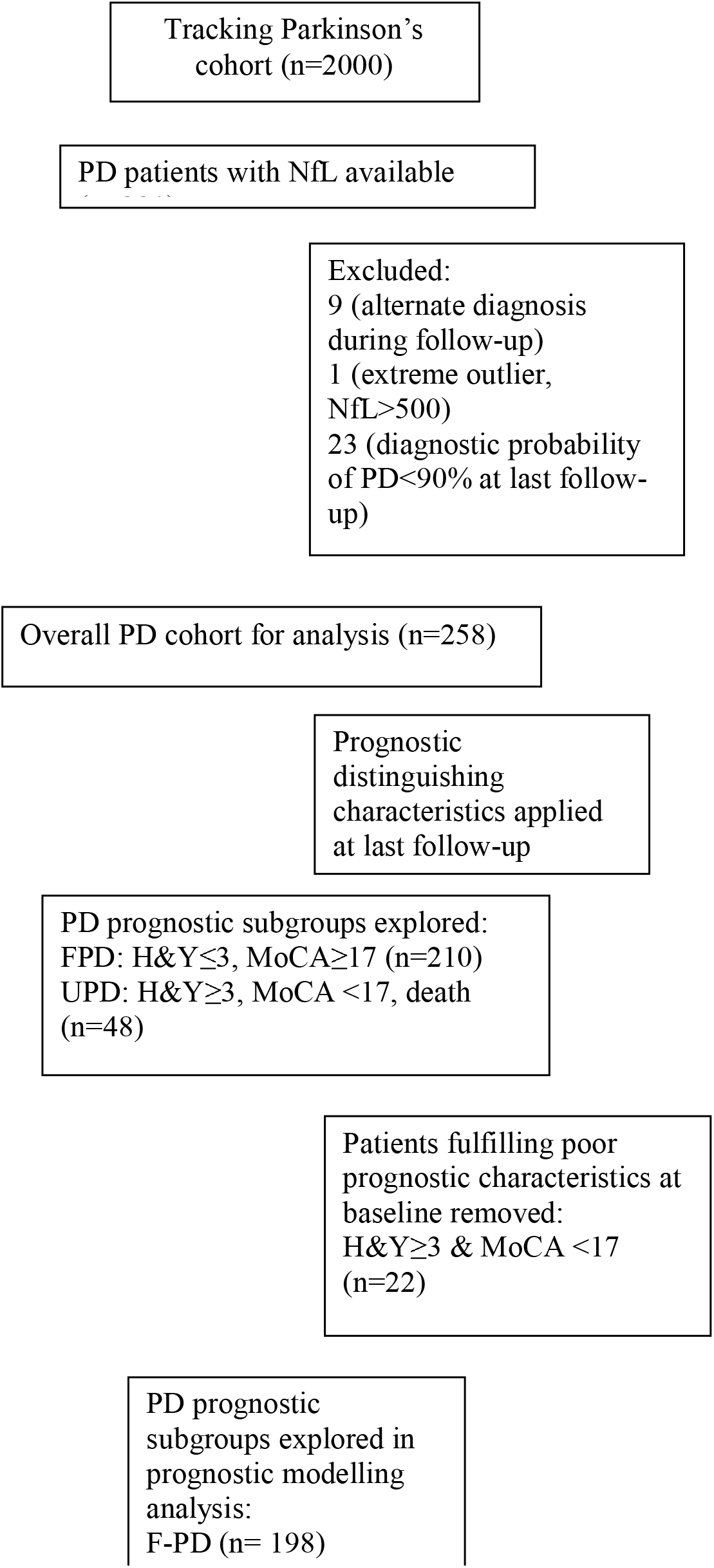
Summary of inclusion and exclusion criteria applied for analysis of cohort and different stages

### Evaluation of NfL as a diagnostic biomarker for PD

Forty-one HC were enrolled in this study (table 1). NfL levels were significantly associated with increasing age but not gender in HC (Coefficient=4.68, P<0.001). NfL was elevated in PD patients at baseline as compared to controls (32.0 ng/L, SD 7.9 vs. 16.4 ng/L, SD 7.5, unadjusted p<0.001, Kruskal-Wallis test). Logistic regression demonstrated a significant difference in NfL levels between HC and PD (Coefficient=2.67, P<0.001), adjusted for age and gender. ROC analysis indicated that serum NfL levels discriminated PD from HC with an AUC of 0.87, 95% CI: 0.82–0.94 (Figure 2b). The optimal cut-off value was 19.4ng/L using the J Youden index with a sensitivity of 77% and specificity of 76%.

**Table 1.**
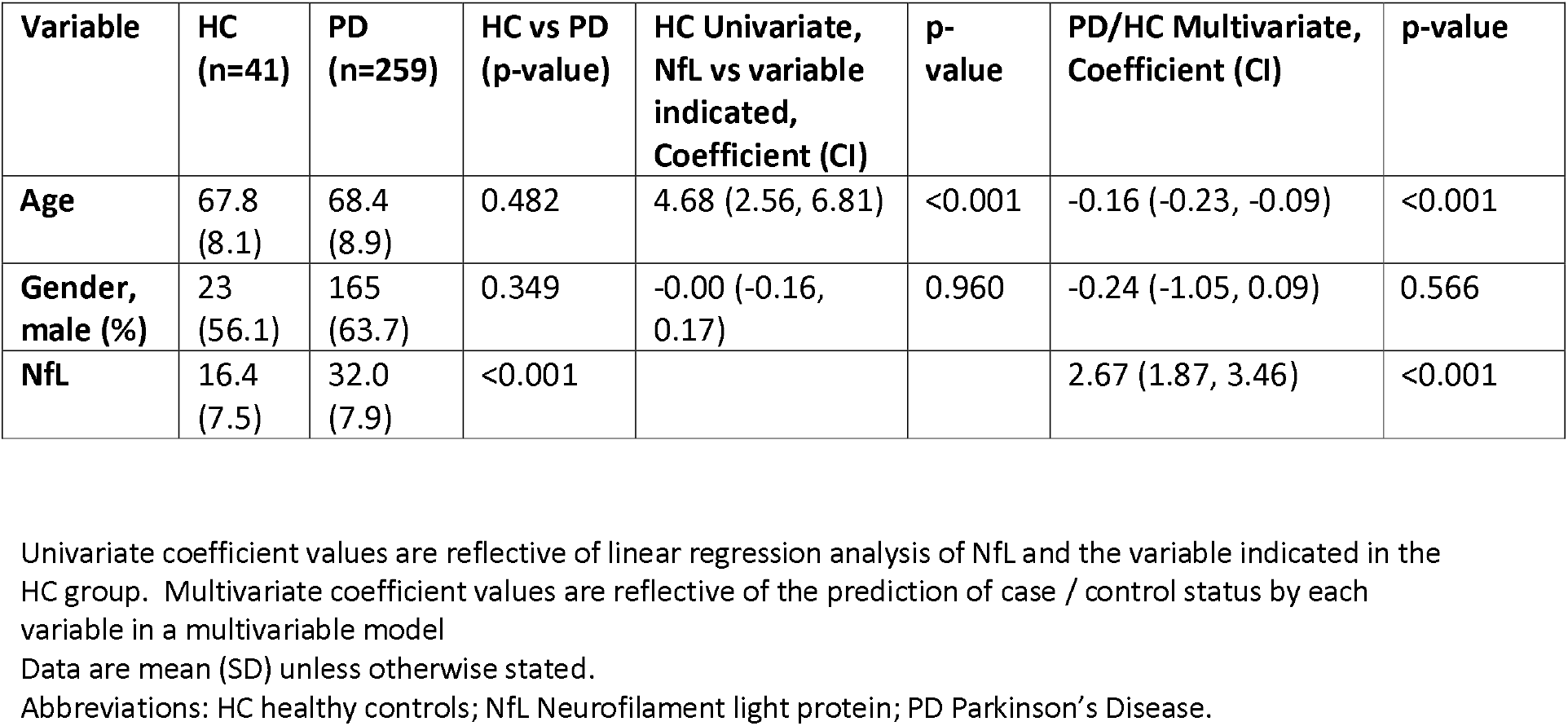
Comparison of healthy control (HC) and Parkinson’s disease (PD) patient characteristics and NfL associations at baseline.

**Figure 2.**
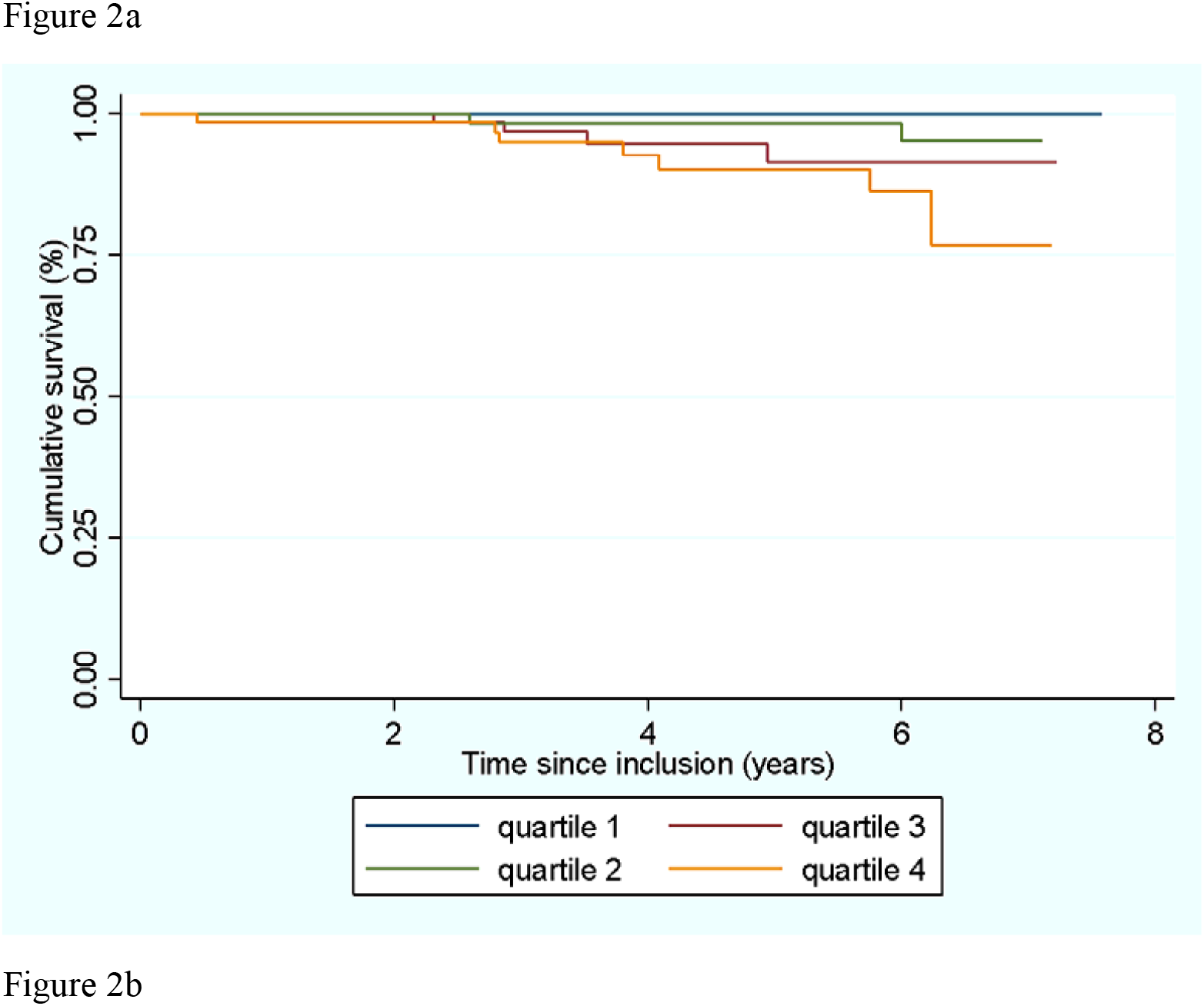
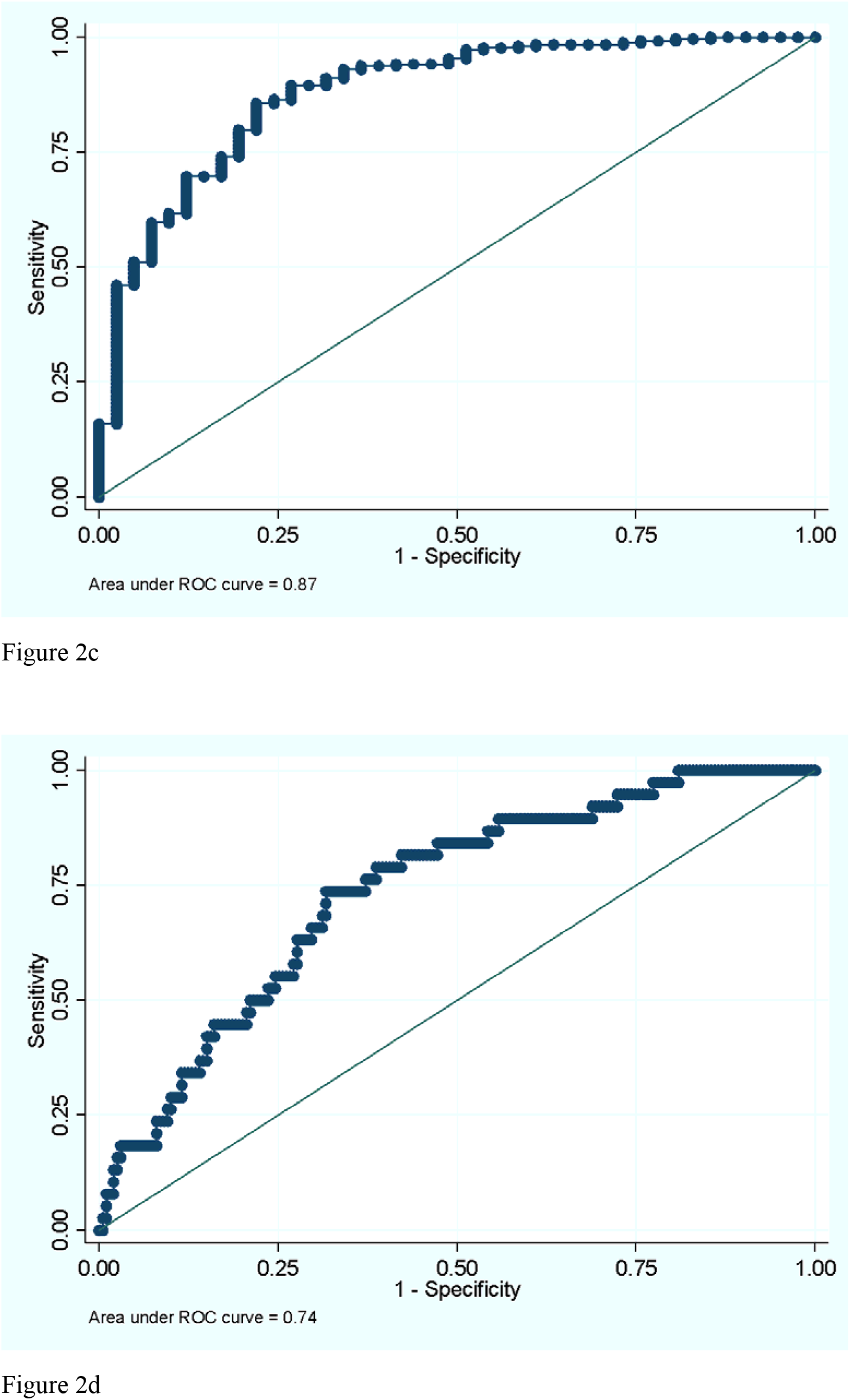
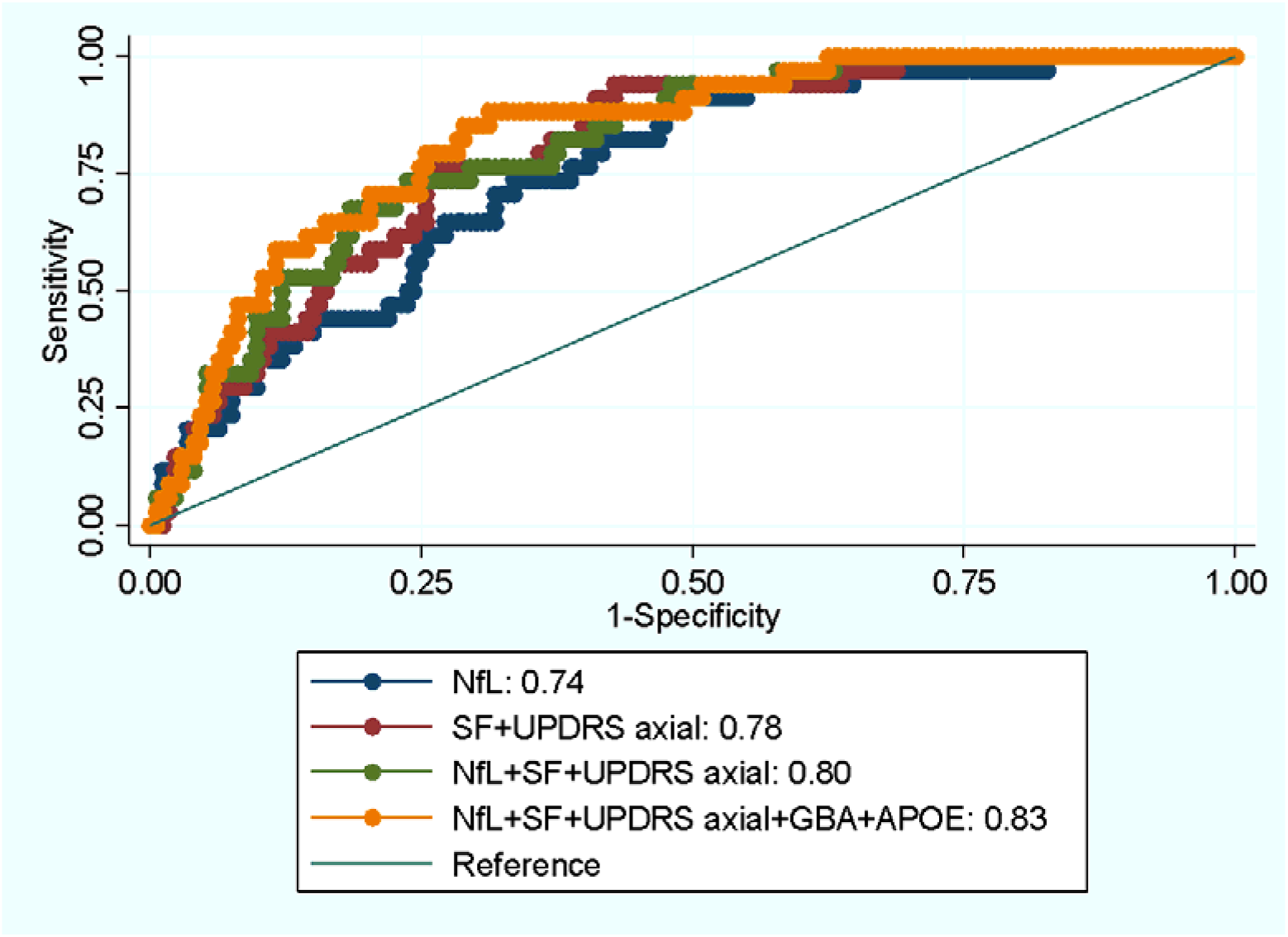
Kaplan-Meir survival estimates and Receiver operator characteristic (ROC) curves Panels: (a) Kaplan-Meir survival estimates of NfL in quartiles (b) ROC curve analysis using NFL to compare HC vs PD, (c) ROC curve analysis using NfL to compare U-PD vs F-PD and (d) combinations of clinical variables and NFL to compare up-PD vs fp-PD. Age & gender were included as covariates in all analysis. Abbreviations fp-PD favourable progression Parkinson’s Disease; UP-PD unfavourable progression Parkinson’s disease.

### Evaluation of the relationship between NFL and clinical features of PD at baseline

PD participant demographics and clinical features at baseline are summarised in Table 2. Serum NfL concentrations were associated with age (Coefficient = 5.86, p <0.001) but not gender or disease duration. Baseline MoCA and semantic fluency (SF) scores were significantly associated with serum NfL levels (MoCA Coefficient -0.60, p = 0.021; SF Coefficient -1.77, p= <0.001), indicating that serum NfL is associated with baseline markers of cognitive impairment. This remained significant for SF after adjustment for age, gender and disease duration. NfL was not associated with measures of functional status at baseline, nor with motor symptom severity measured by the H&Y, MDS-UPDRS 3 total and sub-scores (rigidity, bradykinesia, axial and tremor) (Table 2). No differences in serum NfL levels were noted between patients who carried genetic variants in *APOE* or *GBA* associated with poor prognosis (Supplementary table 2). There was no significant association between NfL levels and *GBA* or *APOE* status. (Table 2)

**Table 2.** Evaluation of the relationship between NFL and clinical features of PD at baseline

| Variables | Mean (SD) or total (%) | Univariate, Coefficient (CI) | P value | Multivariate, Coefficient (CI) | P value |
| --- | --- | --- | --- | --- | --- |
| Age at baseline | 68.4 (8.9) | 5.86 (4.85, 6.86) | <0.001 |  |  |
| Disease duration from diagnosis | 1.3 (0.9) | 0.07 (-0.05, 0.20) | 0.240 |  |  |
| Gender, male (%) | 165 (63.7) | 0.05 (-0.20, 0.13) | 0.692 |  |  |
| *Genetic status* |  |  |  |  |  |
| GBA-positive (non-GD variant) | 18/240 (7.5) | 0.14 (-0.37, 0.65) | 0.590 | 0.30 (-0.12, 0.72) | 0.155 |
| GBA-positive (GD variant) | 10/240 (4.2) | -0.45 (-0.37, 0.65) | 0.590 | 0.02 (-0.51, 0.55) | 0.945 |
| APOE ε4 heterozygous | 63/236 (26.7) | -0.19 (-0.48, 0.09) | 0.186 | 0.09 (-0.14, 0.33) | 0.433 |
| APOE ε4 homozygous | 8/236 (3.4) | 0.34 (-0.37, 1.04) | 0.350 | 0.52 (-0.04, 1.08) | 0.07 |
| *Motor severity outcomes* |  |  |  |  |  |
| H&Y | 1.8 (0.6) | 0.08 (-0.01, 0.16) | 0.068 | 0.01 (-0.09, 0.11) | 0.835 |
| MDS-UPDRS 3 Total | 22.8 (11.6) | -0.73 (-2.37, 0.91) | 0.382 | -1.80 (-3.82, 0.22) | 0.080 |
| MDS-UPDRS rigidity | 3.8 (2.9) | -0.35 (-0.76, 0.05) | 0.085 | -0.43 (-0.92, 0.07) | 0.092 |
| MDS-UPDRS bradykinesia | 10.9 (7.0) | -0.46 (-1.44, 0.52) | 0.354 | -0.80 (-2.01, 0.42) | 0.197 |
| MDS-UPDRS axial | 2.9 (2.6) | 0.34 (-0.03, 0.70) | 0.069 | -0.01 (-0.03, 0.44) | 0.961 |
| MDS-UPDRS tremor | 4.3 (4.0) | -0.37 (-0.94, 0.19) | 0.190 | -0.66 (-1.36, 0.03) | 0.062 |
| *Cognitive Outcomes* |  |  |  |  |  |
| MoCA | 25.1 | -0.60 (-0.04, 0.00) | 0.021 | -0.38 (-1.01, 0.25) | 0.236 |
| Semantic fluency | 21.2 | -1.77 (-2.63, -0.92) | <0.001 | -1.10 (-2.16, 0.04) | 0.043 |
| *Functional outcomes* |  |  |  |  |  |
| SEADL | 86.3 (11.7) | -0.53 (-2.18, 1.11) | 0.524 | 0.57 (-1.48, 2.61) | 0.587 |
| PDQ8 | 6.3 (4.8) | -0.32 (-1.01, 0.36) | 0.353 | 0.41 (-0.41, 1.23) | 0.327 |

Univariate and multivariable (age at baseline, gender and disease duration) linear regression analysis on baseline NfL with baseline clinical measures in PD patients treated as outcome measures. In regression analysis of NfL and genetic status, NfL was treated as the outcome measure and patients who were positive for a genetic mutation were compared to those who were not.
Abbreviations: APO Apolipoprotein; GBA Glucocerebrosidase; H&Y stage Hoehn and Yahr stage; LEDD Levodopa Equivalent Dose; MoCA Montreal Cognitive Assessment; NfL Neurofilament light protein; PD Parkinson's Disease; PDQ8 Parkinson's disease Questionnaire-8; SEADL Schwab and England scale; MDS-UPDRS Movement Disorders Society Unified Parkinson's disease rating scale.

### Association between NFL levels and PD progression and mortality

The MDS-UPDRS score increases with increasing motor impairment, whereas the SEADL decreases with increasing functional impairment. In our analysis of the rate of change of the MDS-UPDRS, a significant negative association with the intercept was noted between baseline NfL and patients overall (total MDS-UPDRS 3 Coefficient -3.55, p=0.001) and sub-section (rigidity, bradykinesia, axial and tremor) motor scores. A similar association was also noted with patients’ overall functional status (SEADL Coefficient 3.36, p=0.004). There was no association between the intercept for cognitive or quality of life scores and NfL. Baseline serum NfL was associated with a more rapid overall progression of motor PD features (as assessed using the total MDS-UPDRS 3, Coefficient 0.79, p=0.012) as well as those thought to be more reflective of underlying disease progression using sub-section motor scores of the UPDRS (UPDRS axial, bradykinesia, rigidity) and the H&Y scores, 0.06, p=0.001 (Table 3). Baseline serum NfL was not significantly associated with the changes in cognition scores (MoCA & SF), though higher levels of NfL at baseline predicted a faster rate of worsening overall function (SEADL Coefficient-1.51, p <0.001).

**Table 3.** Relationship between baseline NfL level and change in motor, cognitive and functional scores using linear mixed effects models

| Variable | Main effect, Coefficient – Intercept (CI) | p-value | Interaction with time - Slope Coefficient (CI) | p-value |
| --- | --- | --- | --- | --- |
| H&Y | -0.11 (-0.23, 0.01) | 0.061 | 0.06 (0.02, 0.08) | 0.001 |
| MDS-UPDRS 3 Total | -3.55 (-5.68, -1.43) | 0.001 | 0.79 (0.17, 1.43) | 0.012 |
| MDS-UPDRS rigidity | -0.82 (-1.38, -0.25) | 0.004 | 0.20 (0.04, 0.36) | 0.016 |
| MDS-UPDRS bradykinesia | -1.87 (-3.14, -0.60) | 0.004 | 0.42 (0.07, 0.77) | 0.019 |
| MDS-UPDRS Axial | -1.20 (-1.94, -0.46) | 0.002 | 0.38 (0.06, 0.70) | 0.018 |
| MDS-UPDRS tremor | -0.79 (-1.54, -0.01) | 0.046 | 0.05 (-0.13, 0.22) | 0.588 |
| MoCA | 0.07 (-0.56, 0.69) | 0.839 | -0.17 (-0.34, 0.01) | 0.062 |
| Semantic fluency | -0.61 (-1.68, 0.46) | 0.263 | -0.03 (-0.31, 0.24) | 0.803 |
| SEADL | 3.36 (1.08, 5.64) | 0.004 | -1.51 (-2.30, -0.72) | <0.001 |
| PDQ8 | 0.02 (-0.86, 0.89) | 0.970 | 0.06 (-0.16, 0.284) | 0.616 |

Model analysis on baseline NfL levels with clinical outcomes in PD patients over time adjusted for age at diagnosis & gender. The main effect indicates the effect of NfL on the intercept and the interaction with time indicates the effect of NfL on the slope (change in value per year) of the model. Abbreviations: CI Confidence Interval; H&Y Hoehn and Yahr stage; MoCA Montreal Cognitive Assessment; NfL Neurofilament light protein; PD Parkinson's Disease; PDQ8 Parkinson's disease Questionnaire-8; SEADL Schwab and England scale; MDS-UPDRS Movement Disorders Society Unified Parkinson's disease rating scale.

Thirteen patients died during follow-up. A higher NfL concentration at baseline predicted a shorter survival, HR 2.40 (1.49-3.88, p<0.001). This remained statistically significant when corrected for age, and gender (HR 1.85, 1.03-3.33, p=0.041). The highest baseline NfL quartile conferred a 2-fold higher risk of mortality in comparison to the lowest quartile (HR 2.04, 1.13-3.69, p=0.018). (Figure 2a)

### Using NFL levels to distinguish an unfavourable prognosis in progression modelling

We explored the value of baseline NfL levels in predicting PD progression within the context of a previously validated prognostic model. We applied distinction criteria (summarised in Figure 1) for determining a poor prognosis at the last available follow-up to separate patients into 2 groups (U-PD & F-PD). PD patients with an unfavourable prognosis (U-PD) had higher serum NfL levels at baseline than those with a favourable prognosis, F-PD (34.9 (SD 18.1) vs 26.4 (SD 13.7), p<0.001). Baseline NfL levels were able to distinguish these phenotypes with an AUC of 0.74, 95% CI 0.66–0.82. (Figure 2c) An optimal cut-off value of 26.7ng/L was determined by the J Youden index with a sensitivity of 67.0% and specificity of 60.0%.

Baseline variables (MDS-UPDRS axial score, SF and NfL) explored in logistic regression individually and in combination with age at the baseline assessment and gender as covariates are summarised in supplementary table 3. The AUC for models incorporating variables individually were SF (0.73, 95% CI 0.65-0.81), MDS-UPDRS axial (0.75, 95% CI 0.67-0.82), and NfL (0.74, 95% CI 0.65-0.82). An AUC of 0.78 (95% CI 0.71-0.85) was seen in the model combining SF and MDS-UPDRS axial scores. The AUC for this model did not significantly differ from the model with NfL alone (0.74 vs 0.78, p=0.22). The addition of NfL to clinical markers did not result in a significant improvement in comparison to clinical markers alone (AUC 0.78 vs 0.80, p=0.30). (Figure 2c) The combination of NfL with both clinical markers did however result in a higher AUC for distinguishing PD progression phenotypes in comparison to NFL alone (0.74 vs 0.80, p=0.02). (Figure 2d) The addition of patient’s combined genetic status and baseline NfL levels into the model resulted in the highest AUC (0.83). This combination resulted in a significantly higher AUC for distinguishing progression phenotypes in comparison to age and gender (0.71 vs 0.83, p=0.0076). (Table 4)

**Table 4.**
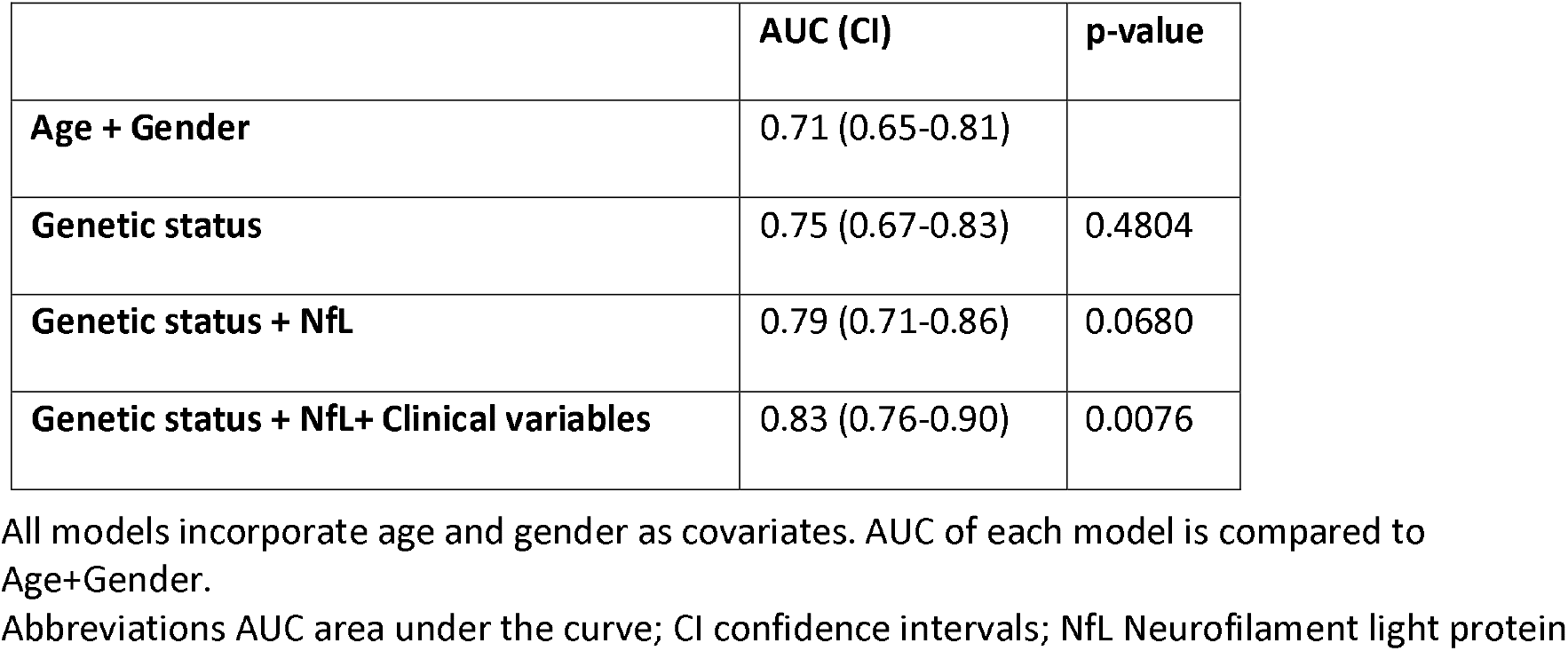
Summary of ROC analysis for models combining baseline predictive variables and comparison of models against model with age and gender

## DISCUSSION

In this study we explored the use of serum NfL as a potential prognostic biomarker in a large and well-studied cohort of recently diagnosed PD patients with prolonged follow-up and high clinical diagnostic certainty. We found baseline NfL to be associated with age and aspects of cognition while also being an acceptable marker for distinguishing PD from controls. We have also established that serum NfL can predict several aspects of PD progression while also providing additional prognostic value when combined with previously validated clinical measures in prognostic modelling.

Serum NfL is higher in older PD patients and unaffected controls. This presumably relates to increased axonal degeneration and decreased clearance that occurs with ageing.[21–23] If NfL is used as a diagnostic and/or prognostic tool then age adjusted/corrected measures will be required.

Serum NfL levels were elevated in PD compared to HC. This finding has been inconsistent in prior studies [23–26]. Potential reasons for these variable findings include underpowered sample sizes and variability in selection criteria for control groups [10,27]. Our enrolment of HC without a history of neurological disorders potentially explains the significant difference noted in comparison to PD. A further potential explanation for this discrepancy could be our measurement of NfL in the earlier stages of symptomatic PD (mean disease duration of 1.3 years), since NfL levels appear to peak in the stages of conversion to clinically manifest PD before gradually declining [28,29]. Taken together these findings support the potential use of NfL as a biomarker for clinical trial recruitment in early PD.

We did not find an association between baseline motor severity measures (MDS-UPDRS-3 & H&Y) though a trend towards significance was noted. The significance of association between the MDS-UPDRS 3 (total and sub-scores) and NfL has varied between studies. A potential explanation for this could be the discrepant use of ‘ON’, ‘OFF’ and treatment naive UPDRS scores. In our study most patients were assessed in the ON state thus making correction for this of limited value. The association of H&Y status and NfL appears to be more consistent in studies [13,14,24,30]. This is potentially attributed to the H&Y stages more prominently reflecting the patient’s axial status at higher levels (>2.5) which seems to better correlate with NfL while also being related to reduction in white matter integrity in the substantia nigra [31]. The lack of significant association between H&Y and NfL at baseline in our cohort is likely a reflection of the minimal representation of patients with more severe H&Y scores at this assessment time point.

We found that baseline MoCA and semantic fluency scores were inversely associated with NfL levels. This finding is consistent with other studies exploring global cognitive function. The clearer association between sematic fluency and NfL noted is intuitive and consistent with a previous study that explored this particular subdomain in the MMSE [32]. A deficit in this test is a reflection of fronto-temporal dysfunction.[33] Abnormalities in axonal tracts in these regions have been noted in the early stages of PD and seem to correlate with CSF NfL levels [14]. This finding potentially highlights the value of more detailed neuropsychological testing, but this is of course more labour intensive than a simple blood test.

Despite a previous study suggesting higher blood NfL levels in patients with more pathogenic variants of *GBA* [23], we did not replicate this finding. Furthermore, we did not note significant differences in NfL levels when comparing patients with a heterozygous or homozygous APOE ε4 status to those who did not. These genetic markers are of interest considering their variable association with more severe cognitive and motor progression [7,34]. The interpretation of these findings are however mitigated by the small number of patients in this cohort with these genetic findings and future collaborative studies exploring their interaction with NfL levels would be of interest. We have previously found that baseline cognition in the PROBAND cohort is not associated with GBA mutation status. These data could be interpreted as meaning that GBA and ApoE are associated with progression of pathology after diagnosis, as compared to NFL which reflects the intensity of neurodegeneration at baseline.

We found that serum NfL levels could predict progression of motor, and functional status while also predicting mortality in PD. We noted a negative main effect of higher baseline NfL levels on progression scores in mixed modelling. This novel finding is potentially consistent with NfL levels peaking prior to the onset of appreciable clinical features [28]. Our observation of higher baseline NfL levels predicting more rapid motor and functional progression mirrors several other studies [12–14,29,30] and could potentially be explained by NfL’s ability to reflect baseline pathological characteristics which predict a more malignant progression such as the magnitude of alpha synuclein deposition and anatomical dysfunction present [35–37]. The potential for a regression towards the mean phenomenon partly explaining our observation does however need to be acknowledged. Our finding of baseline serum NfL predicting functional progression as measured by the SEADL scale has not previously been reported, and may reflect the relationship between disease burden and disability/function [38]. The lack of association noted with cognitive score progression is out of keeping with previous studies and may reflect the limited number of patients who progressed to develop significant cognitive dysfunction (dementia) in our cohort. NfL appears to better predict this than changes confined to ranges in normal or mild cognitive impairment [30].

PD progression and prognosis can be highly variable. A number of phenotypes have previously been explored with the goal of predicting future outcomes[39]. To date, studies focusing on the potential role of NfL in predicting more severe progression phenotypes have suggested that patients with a more prominent postural disability phenotype have more substantial increases in NfL levels over time [13,14]. Our goal was to explore if NfL levels could play a role in a model which predicts PD progression in a more encompassing and practical manner that could potentially be utilised in disease modifying clinical trials. In this regard we explored if baseline NfL could replace or complement a number of simple clinical markers previously identified to predict PD progression in a well validated model[40]. While we did not find that NfL provided significant additional value to the clinical variables previously identified, we did note that it could replace the combination of these markers without compromising modelling accuracy. This finding could support its future use in randomising patients between active treatment and placebo arms in clinical trials. While our findings suggest an encouraging role for NFL in a future prognostic predictive capacity, its value was strongest when combined with clinical variables and patient’s genetic status. This finding highlights the importance of combining biomarkers with clinical scales though the addition of a more specific biomarker would likely be necessary to improve progression modelling more significantly.

The strengths of our study are its large sample size and prolonged follow-up of up to 72 months. We were limited by a lack of assessment in the ‘OFF’ medication state which restricts our ability to interpret NfL associations with overall motor progression and therefore its value in clinical trial modelling where MDS-UPDRS OFF state changes may be the primary outcome. We also lack neuropathological diagnostic confirmation in our cohort although our exclusion of patients with a diagnostic probability of <90% at the last available visit aimed to mitigate the potential inclusion of misdiagnosed patients.

We were able to demonstrate that serum NfL is a useful biomarker for prediction of PD progression. In the appropriate setting, NfL could potentially be used to enrich a clinical trial cohort for individuals likely to have more rapid disease progression, which might then shorten the follow up time required to detect a disease modifying signal, or alternatively to help ensure that randomised groups are more likely to be balanced in terms of progression rates, thus facilitating detection of agents with true disease modifying properties.

## Data Availability

All data produced in the present study are available upon reasonable request to the authors

## Cohort studies

Tracking Parkinson’s is primarily funded and supported by Parkinson’s UK. It is also supported by the National Institute for Health Research (NIHR) Dementias and Neurodegenerative Diseases Research Network (DeNDRoN). This research was supported by the National Institute for Health Research University College London Hospitals Biomedical Research Centre and Cambridge BRC. The UCL Movement Disorders Centre is supported by the Edmond J. Safra Philanthropic Foundation.

## Genetic and biomarker analysis

Work on the genetics and biomarkers of progression in Parkinson’s and related disorders is supported by Parkinson’s UK (PhD Studentship to Dr Tan H-1703, Understanding and predicting Parkinson’s progression), and the PSP Association. The study is funded by the joint efforts of The Michael J. Fox Foundation for Parkinson’s Research (MJFF) and the Aligning Science Across Parkinson’s (ASAP) initiative. MJFF administers the grant [Grant ID: ASAP-000478] on behalf of ASAP and itself. For the purpose of open access, the author has applied a CC-BY public copyright license to the Author Accepted Manuscript (AAM) version arising from this submission.

## Competing Interests

NV has received unconditional educational grants from IPSEN and Biogen, travel grants from IPSEN, AbbVie and The International Parkinson’s Disease and Movement Disorders Society, speaker’s honorarium from AbbVie and STADA and served on advisory boards for Abbvie and Brittania outside of the submitted work.

ML No competing interest

AH No competing interest

TG No competing interest

MT No competing interest

EJ No competing interest

RR No competing interest

JW No competing interest

KAG No competing interest

VC No competing interest

DA No competing interest

CG No competing interest

RAB receives consultancy monies from Novo Nordisk; UCB; BlueRock therapeutics; Aspen Neuroscience and FCDI. He also receives grant support from the MRC, Wellcome, ASAP, EU, NIHR, Cure Parkinson’s Trust, John Black Charitable Foundation, PUK, and Rosetrees Trust. He receives royalties from Wiley and Springer Nature.

JH No competing interest

NW No competing interest

HH No competing interest

NMW No competing interest

YB-S No competing interest

HZ has served at scientific advisory boards for Abbvie, Alector, Eisai, Denali, Roche Diagnostics, Wave, Samumed, Siemens Healthineers, Pinteon Therapeutics, Nervgen, AZTherapies and CogRx, has given lectures in symposia sponsored by Cellectricon, Fujirebio, Alzecure and Biogen, and is a co-founder of Brain Biomarker Solutions in Gothenburg AB (BBS), which is a part of the GU Ventures Incubator Program (outside submitted work). DG has received honoraria from BIAL Pharma, GE Healthcare, and Vectura plc, and consultancy fees from the Glasgow Memory Clinic.

TF has received grants from National Institute of Health Research, Michael J Fox Foundation, John Black Charitable Foundation, Cure Parkinson’s Trust, Innovate UK, Van Andel Research Institute and Defeat MSA. He has served on Advisory Boards for Voyager Therapeutics, Handl therapeutics, Living Cell Technologies, Bial, Profie Pharma. He has received honoraria for talks sponsored by Bial, Profile Pharma, Boston Scientific.

HRM is employed by UCL. In the last 24 months he reports paid consultancy from Biogen, UCB, Abbvie, Denali, Biohaven, Lundbeck; lecture fees/honoraria from Biogen, UCB, C4X Discovery, GE-Healthcare, Wellcome Trust, Movement Disorders Society; Research Grants from ASAP, Parkinson’s UK, Cure Parkinson’s Trust, PSP Association, CBD Solutions, Drake Foundation, Medical Research Council. Dr Morris is a co-applicant on a patent application related to C9ORF72 - Method for diagnosing a neurodegenerative disease (PCT/GB2012/052140)

## Funding

HZ is a Wallenberg Scholar supported by grants from the Swedish Research Council (#2018-02532), the European Research Council (#681712), Swedish State Support for Clinical Research (#ALFGBG-720931), the Alzheimer Drug Discovery Foundation (ADDF), USA (#201809-2016862), the AD Strategic Fund and the Alzheimer’s Association (#ADSF-21-831376-C, #ADSF-21-831381-C and #ADSF-21-831377-C), the Olav Thon Foundation, the Erling-Persson Family Foundation, Stiftelsen för Gamla Tjänarinnor, Hjärnfonden, Sweden (#FO2019-0228), the European Union’s Horizon 2020 research and innovation programme under the Marie Skłodowska-Curie grant agreement No 860197 (MIRIADE), and the UK Dementia Research Institute at UCL.

DG has received grant funding from the Neurosciences Foundation, Michael’s Movers, and Parkinson’s UK.

## Supplementary Tables

**Supplementary Table 1.**
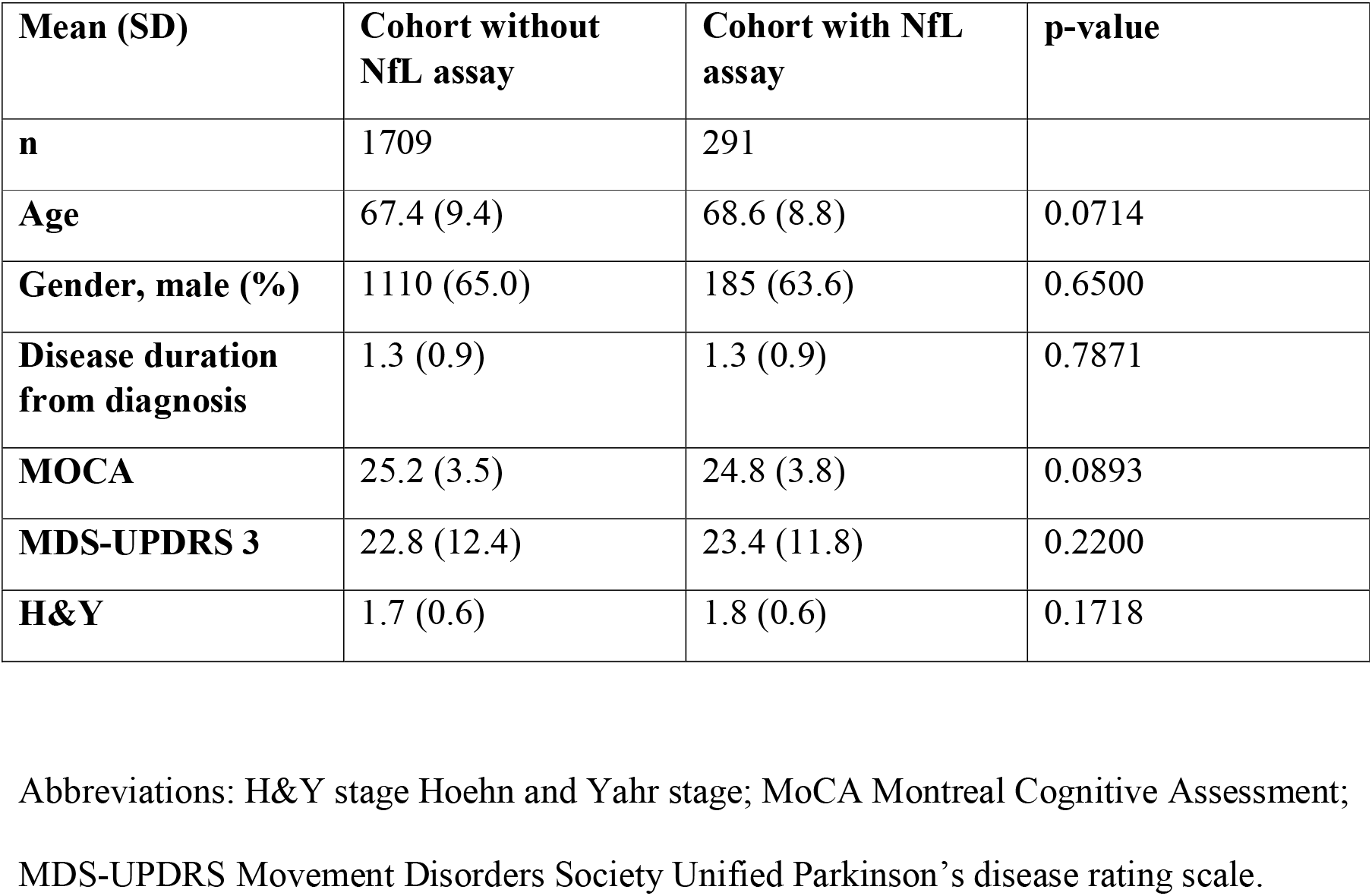
Baseline characteristics of patients selected for NfL analysis compared to remaining cohort

| **Mean (SD)** | **Cohort without NfL assay** | **Cohort with NfL assay** | **p-value** |
| --- | --- | --- | --- |
| **n** | 1709 | 291 |  |
| **Age** | 67.4 (9.4) | 68.6 (8.8) | 0.0714 |
| **Gender, male (%)** | 1110 (65.0) | 185 (63.6) | 0.6500 |
| **Disease duration from diagnosis** | 1.3 (0.9) | 1.3 (0.9) | 0.7871 |
| **MOCA** | 25.2 (3.5) | 24.8 (3.8) | 0.0893 |
| **MDS-UPDRS 3** | 22.8 (12.4) | 23.4 (11.8) | 0.2200 |
| **H&Y** | 1.7 (0.6) | 1.8 (0.6) | 0.1718 |

Abbreviations: H&Y stage Hoehn and Yahr stage; MoCA Montreal Cognitive Assessment;
MDS-UPDRS Movement Disorders Society Unified Parkinson's disease rating scale.

**Supplementary table 2.** Comparison of baseline characteristics of patients with and without genetic abnormalities

| Mean (SD) | GBA negative PD (n=213) | Non-GD variant PD (n=17) | GD variant PD (n=10) | Non ε4 allele PD (n=165) | Heterozygous ε4 PD (n=63) | Homozygous ε4 PD (n=8) |
| --- | --- | --- | --- | --- | --- | --- |
| Age | 68.6 (8.8) | 67.6 (8.3) | 62.0 (11.2) | 69.7 (8.7) | 65.4 (8.2) ** | 67.5 (4.9) |
| Gender, male (%) | 132 (62.0) | 15 (88.2) | 6 (60.0) | 112 (67.9) | 36 (57.1) | 5 (62.5) |
| Disease duration | 1.3 (0.9) | 1.1 (0.7) | 1.1 (1.1) | 1.3 (0.9) | 1.3 (0.9) | 1.0 (0.7) |
| H&Y | 1.8 (0.6) | 1.8 (0.6) | 1.7 (0.7) | 1.8 (0.6) | 1.7 (0.6) | 1.4 (0.6) |
| MDS-UPDRS 3 | 23.2 (11.5) | 20.1 (12.6) | 19.7 (16.8) | 23.0 (11.8) | 23.1 (10.5) | 15.9 (10.4) |
| MOCA | 25.1 (3.6) | 25.7 (2.3) | 24.9 (2.8) | 25.2 (3.3) | 24.9 (3.4) | 24.0 (2.9) |
| SF | 20.9 (6.3) | 22.8 (6.6) | 26.2* (4.2) | 21.4 (6.3) | 21.2 (6.4) | 18.5 (10.4) |
| NfL | 30.5 (17.3) | 32.2 (17.4) | 25.8 (17.0) | 30.6 (17.2) | 27.4 (13.2) | 36.7 (22.9) |

\* GD variant PD vs GBA negative PD $p < 0.05$ , \*\* Heterozygous ε4 PD vs Non ε4 allele PD $p < 0.01$
Abbreviations: GD Gaucher disease; H&Y stage Hoehn and Yahr stage; MoCA Montreal Cognitive Assessment; NfL Neurofilament light protein; PD Parkinson's Disease; MDS-UPDRS Movement Disorders Society Unified Parkinson's disease rating scale; SF Semantic fluency.

**Supplementary table 3.** Regression coefficients of the final combination models explored.

| Model 1 | Coefficient | Standard error | P value |
| --- | --- | --- | --- |
| Intercept | -4.52 | 1.96 |  |
| Age | 0.04 | 0.03 | 0.115 |
| Gender | -0.96 | 0.46 | 0.030 |
| NfL | 0.68 | 0.23 | 0.003 |
| Model 2 |  |  |  |
| Intercept | -5.20 | 1.98 |  |
| Patient age | 0.07 | 0.03 | 0.009 |
| Gender | -0.69 | 0.45 | 0.123 |
| UPDRS axial | 0.23 | 0.09 | 0.008 |
| Semantic fluency | -0.07 | 0.03 | 0.030 |
| Model 3 |  |  |  |
| Intercept | -6.74 | 2.14 |  |
| Age | 0.03 | 0.03 | 0.271 |
| Gender | -0.96 | 0.48 | 0.046 |
| UPDRS axial | 0.26 | 0.09 | 0.004 |
| Semantic fluency | -0.06 | 0.03 | 0.097 |
| NfL concentration | 0.54 | 0.23 | 0.016 |
| Model 4 |  |  |  |
| Intercept | -5.49 | 2.63 |  |
| Age | 0.06 | 0.03 | 0.099 |
| Gender | -1.26 | 0.57 | 0.028 |
| UPDRS Axial | 0.29 | 0.10 | 0.004 |
| Sematic Fluency | -0.06 | 0.04 | 0.131 |
| NfL concentration | 0.58 | 0.27 | 0.030 |
| ApoE status | 0.77 | 0.37 | 0.036 |
| GBA status | 0.47 | 0.49 | 0.341 |

**Supplementary table 4.** Summary of ROC analysis for models using different baseline predictive variables and comparison of models

|  | AUC | CI |
| --- | --- | --- |
| Age+gender | 0.71 | 0.63-0.79 |
| 1. NfL | 0.74 | 0.65-0.82 |
| 2. UPDRS Axial | 0.75 | 0.67-0.82 |
| 3. SF | 0.73 | 0.65-0.81 |
| 4. GBA status | 0.73 | 0.64-0.81 |
| 5. APOE status | 0.73 | 0.64-0.82 |
| 6. UPDRS Axial/NfL | 0.77 | 0.70-0.85 |
| 7. SF/NFL | 0.76 | 0.68-0.84 |
| 8.GBA/NFL | 0.76 | 0.68-0.84 |
| 9.ApoE/NfL | 0.76 | 0.68-0.84 |
| 10. SF/UPDRS Axial | 0.78 | 0.71-0.85 |
| 11. SF/UPDRS Axial/NfL | 0.80 | 0.73-0.87 |
| 12. SF/UPDRS Axial/GBA | 0.80 | 0.72-0.87 |
| 13. SF/UPDRS Axial/apoE | 0.79 | 0.71-0.86 |
| 14. SF/UPDRS<br>Axial/NfL/GBA status | 0.81 | 0.73-0.88 |
| 15. SF/UPDRS<br>Axial/NfL/ApoE | 0.81 | 0.73-0.88 |
| 16. SF/UPDRS<br>Axial/NfL/ApoE/GBA | 0.83 | 0.76-0.90 |
| AUC comparison | Chi | p-value |
| 1 vs 10 | 1.48 | 0.22 |
| 1 vs 11 | 5.18 | 0.02* |
| 1 vs 14 | 4.73 | 0.03* |
| 1 vs 15 | 5.68 | 0.02* |
| 1 vs 16 | 5.40 | 0.02* |
| 10 vs 14 | 0.58 | 0.45 |
| 10 vs 15 | 2.06 | 0.15 |
| 10 vs 16 | 2.16 | 0.14 |

All models incorporate age and gender as covariates
Abbreviations AUC area under the curve; NfL Neurofilament light protein; SF semantic fluency; UPDRS Unified Parkinson's Disease Rating Scale total axial score; SF semantic fluency

## Notes

### Competing Interest Statement

The authors have declared no competing interest.

### Clinical Protocols

https://dx.doi.org/10.17504/protocols.io.bzbep2je

https://dx.doi.org/10.17504/protocols.io.by9ypz7w

https://dx.doi.org/10.17504/protocols.io.bzd7p29n

### Funding Statement

Cohort studies: Tracking Parkinsons is primarily funded and supported by Parkinsons UK. It is also supported by the National Institute for Health Research (NIHR) Dementias and Neurodegenerative Diseases Research Network (DeNDRoN). This research was supported by the National Institute for Health Research University College London Hospitals Biomedical Research Centre and Cambridge BRC. The UCL Movement Disorders Centre is supported by the Edmond J. Safra Philanthropic Foundation.
Genetic and biomarker analysis: Work on the genetics and biomarkers of progression in Parkinsons and related disorders is supported by Parkinsons UK (PhD Studentship to Dr Tan H-1703 Understanding and predicting Parkinsons progression) and the PSP Association. The study is funded by the joint efforts of The Michael J. Fox Foundation for Parkinsons Research (MJFF) and the Aligning Science Across Parkinsons (ASAP) initiative. MJFF administers the grant [Grant ID: ASAP-000478] on behalf of ASAP and itself. For the purpose of open access the author has applied a CC-BY public copyright license to the Author Accepted Manuscript (AAM) version arising from this submission.

### Author Declarations

Tracking Parkinsons study (REC Reference: 11/AL/0163) and PROSPECT (REC Reference: 14/LO/1575) studies have multicentre research ethics committee approvals

